# Prostate cancer castrate resistant progression usage of non-canonical androgen receptor signaling and ketone body fuel

**DOI:** 10.1101/2021.01.12.21249395

**Authors:** Estefania Labanca, Juan Bizzotto, Pablo Sanchis, Jun Yang, Peter D.A. Shepherd, Alejandra Paez, Valeria Antico-Arciuch, Nicolas Anselmino, Sofia Lage-Vickers, Anh G. Hoang, Mark Titus, Eleni Efstathiou, Javier Cotignola, John Araujo, Christopher Logothetis, Elba Vazquez, Nora Navone, Geraldine Gueron

## Abstract

Prostate cancer (PCa) that progresses after androgen deprivation therapy (ADT) remains incurable. The intricacy of metabolic pathways associated with PCa progression spurred us to develop a metabolism-centric analysis. Using PCa patient-derived xenografts (PDXs) we assessed the metabolic changes after castration of tumor-bearing mice. We found that relapsed tumors had a significant increase in fatty acids and ketone body content compared with baseline. We confirmed that critical ketogenic/ketolytic enzymes (ACAT1, OXCT1, BDH1) were significantly augmented after castrate-resistant progression. Further, these enzymes are increased in the human donor tissue after progressing to ADT.

Increased ACAT1 and OXCT1 was also observed for a subset of PCa patients that relapsed with low AR and ERG expression. These factors were associated with decreased biochemical relapse and progression free survival. In summary, our studies reveal the key metabolites fueling castration resistant progression in the context of a partial or complete loss of AR dependence.

## INTRODUCTION

Androgen signaling inhibition remains the mainstay, and patients are treated similarly despite the known heterogeneity of responsiveness. The underlying mechanisms that account for the ultimate emergence of resistance to androgen deprivation therapy (ADT), progressing to castrate-resistant prostate cancer (CRPC), and to second generation androgen blockade, include those that reactivate androgen receptor (AR) signaling despite strong inhibition (e.g., AR amplification and/or mutation), or those that are entirely independent or cooperate with androgen signaling to underlie PCa progression (1).

It is well known that oncogenic alterations modify the metabolic program of cancer cells (2). Mammalian cells use two main pathways to generate energy in the form of adenosine triphosphate (ATP) from glucose: oxidative phosphorylation in the mitochondria, with CO_2_ and H_2_O as final products, and glycolysis in the cytoplasm, yielding lactate. It is known that during neoplastic transformation, glucose is metabolized to lactate even in the presence of oxygen, the Warburg effect (3, 4).

The hypoxic conditions commonly observed in tumors, force cancer cells to adapt and to exploit alternative fuel sources (5, 6). In PCa, the metabolic pathways of greatest relevance in this context seem to be fatty acid (FA) and glutamine metabolism (7). In a meta-analysis of clinical metabolic profiling of 136 cancer cohorts including blood, tissue, and urine samples, ketone bodies (KB) (known products of FA metabolism), and in particular β-hydroxybutyrate (BHBA), were identified among the most upregulated metabolites compared with normal specimens (8). KB metabolism provides substrate availability for lipogenesis, critical for tumor biomass expansion. Further, gain of this lipogenic conduit is associated with shorter patient survival and greater tumor burden (9). Expression of mitochondrial acetyl-CoA acetyltransferase (ACAT1) has been associated with aggressive PCa and biochemical recurrence after ADT (10), indicating that ketogenesis/ketolysis may also play a role in PCa progression.

The intricacy of energy metabolism pathways associated with neoplastic transformation and cancer progression is partially responsible for the extensive gaps in our understanding of this phenotype, with an obvious negative impact on our ability to harness them for clinical purposes. Progress in comprehending changes in the metabolic program during PCa progression has also been hampered by a lack of models, representative of the clinical spectrum and biologic complexity of PCa. Patient-derived xenografts (PDXs) have been developed and have led to therapeutically relevant approaches (11-14). It is imperative to harness these available tools to dissect how this metabolic plasticity orchestrates progression and metastasis.

In this work, using PDXs that mimic the response of the human donor to ADT in a well-controlled study, we performed for the first time a comprehensive metabolomic analysis of PCa PDXs that relapsed following castration. We discovered a metabolic shift from high glycolytic activity to exacerbated KB metabolism, indicating that a subpopulation of CRPCs that progress with partial or complete loss of AR dependence are fueled by KB. We confirmed that expression of critical ketogenic/ketolytic enzymes is significantly augmented after castration-resistant progression in both the PDX tumor and its human donor tissue. Further, we assessed the expression of these ketogenic/ketolytic enzymes in a subset of PCa patients that relapse with low androgen receptor (AR) and ETS-related gene (ERG) and also analyzed the correlation with biochemical relapse and progression free survival.

## RESULTS

### PDXs mimic short- and long-term response to ADT in human donor

To model the response of PCa to castration we used a PCa PDX, MDA PCa 183, derived from a bone metastasis in a treatment naïve patient. A genomic characterization previously performed in the MDA PCa 183 tumors (whole genome, exome, and transcriptome sequencing) identified *PTEN* homozygous deletion, *TMPRSS2/ERG* and *TP53*/*SCFD1* rearrangements, and *PLK1* and *ERG* outlier expressions (15). Furthermore, this PDX expresses wild-type AR and ERG, as assessed by IHC (15, 16).

We subjected mice bearing MDA PCa 183 tumors to surgical castration (**Fig 1 A**) and found a significant reduction in tumor volume with a concomitant drop in PSA blood levels (**Fig 1 B**). An analysis of tumor volume before and after castration demonstrated that there are statistically significant differences in the tumor growth slopes in intact vs castrated mice (17), In parallel, the response of the patient, donor of MDA PCa 183, to a gonadotropin-releasing hormone (GnRH) antagonist (**Fig 1 A**) was reflected by a drop in PSA blood levels (**Fig 1 B**) (similar to the findings in MDA PCa 183 after castration), accompanied by improvement of symptoms associated with tumor burden.

**Fig 1.**
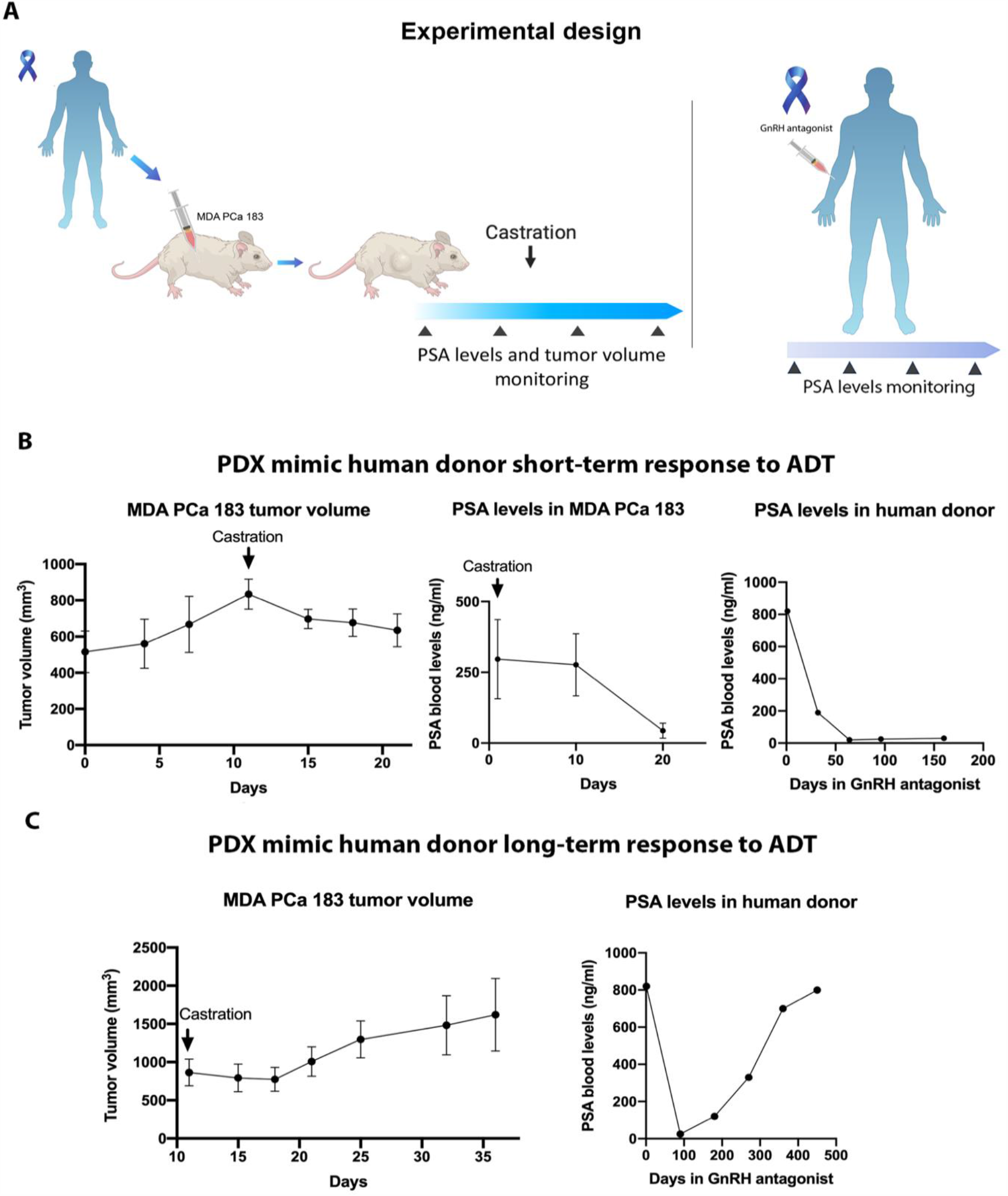
PDXs mimic response to ADT. (A) Schematic representation of subcutaneous injection of MDA PCa 183 (PCa bone metastasis) in CB17 SCID mice followed by castration when tumors reached a volume of 500 mm^3^ or higher (left panel) and human donor of MDA PCa 183 treated with ADT using GnRH antagonist (right panel). (B) and (C) Tumor volume and PSA levels of MDA PCa 183 subcutaneous tumors in mice and PSA levels assessed in response to androgen ablation in human donor of MDA PCa 183. Data are represented as mean ± SD.

In a long-term follow up we observed that a proportion of MDA PCa 183 tumors in castrated mice relapsed (**Fig 1 C**). Similarly, the patient response after 4 months undergoing ADT had evidence of disease progression according to an assessment of PSA levels (**Fig 1 C**), worsening of bone scan findings and bone metastasis-associated pain.

### Modeling and characterization of PCa response to castration

To model PCa response to castration we obtained MDA PCa 183 tumors growing in intact mice (Control), those harvested 10 days after mice castration (when serum testosterone reached castration levels; early response to castration (8)) and at relapse (Relapse) (**Fig 2 A**). As depicted in **Fig 2 B**, MDA PCa 183 shows an adenocarcinoma tumor densely packed before castration (Control). At ERC and Relapse tumors seem less dense.

**Fig 2.**
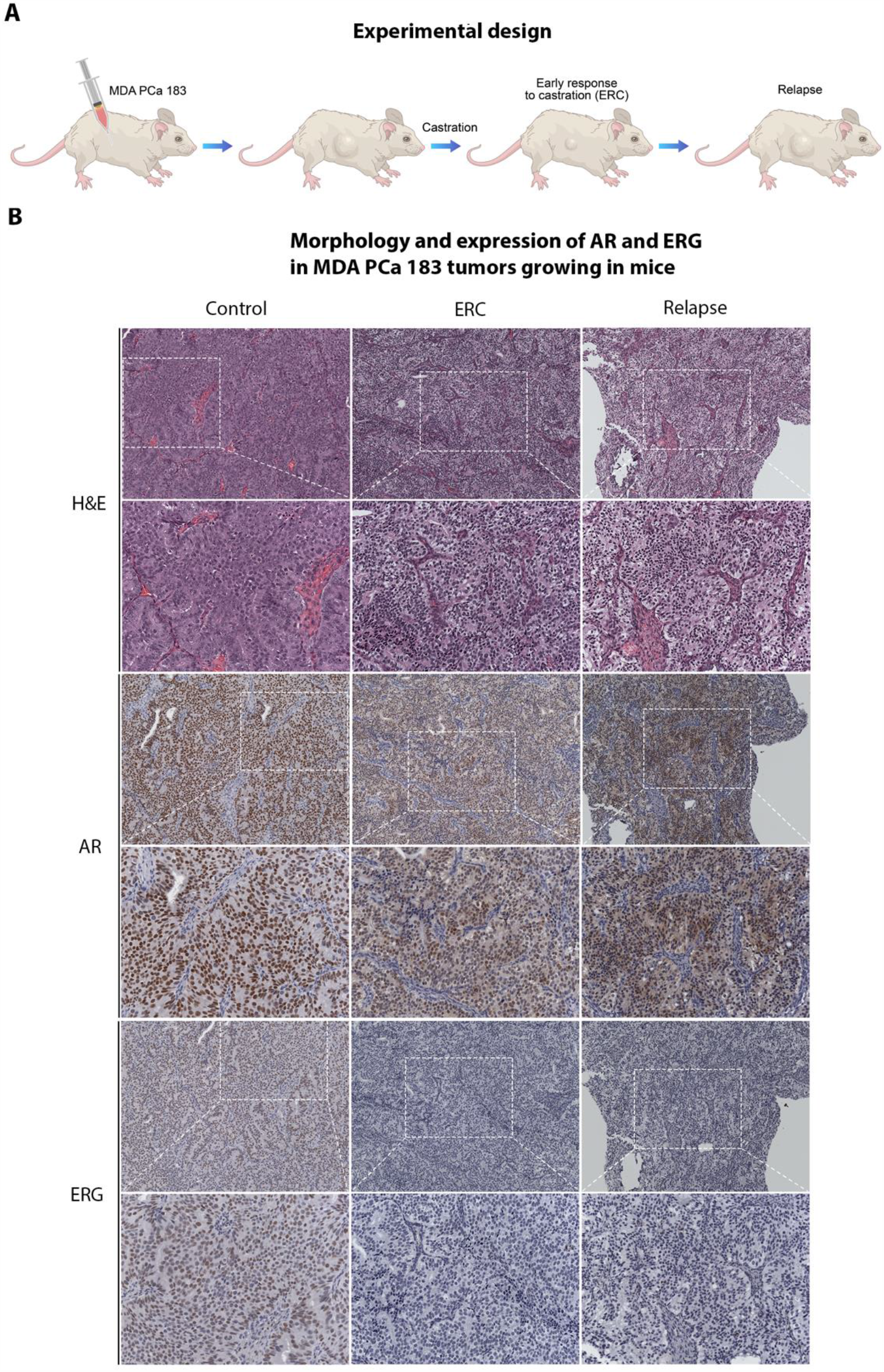
AR and ERG expression in MDA PCa 183 tumors in progression to castration. (A) Schematic representation of response to castration in mice bearing MDA PCa 183 tumor. (B) Representative photomicrograph images of MDA PCa 183 tumors (Control [n=5], ERC [n=6], and Relapse [n=3]) sections immunostained with AR and ERG. (Magnification 200X) ERC: early response to castration; AR: androgen receptor; ERG: ETS-related gene.

IHC analysis revealed that MDA PCa 183 tumors growing in control mice expressed high nuclear AR in nearly all cells, no cytoplasmic staining was observed. ERG staining was present in the nuclei of all cells (**Fig 2 B**). At ERC, where tumor growth was halted, AR expression was lower, with heterogeneous nuclear localization and scarce cytoplasmic staining; while ERG reactivity was negative (**Fig 2 B**). Likewise, in relapsed tumors AR expression was lower with cytoplasmic and nuclear localization and ERG staining remained negative (**Fig 2 B**). These results suggest that MDA PCa 183 progressed to castration independently of AR signaling or by activating non-canonical AR signals.

We furthered our study performing an RNA-Seq analysis of 37 CRPC MDA PCa PDXs, (16, 18) focusing on AR and AR downstream targets’ gene expression (*ERG, TMPRSS2, KLK3, CAMKK2, NKX3*.*1, FKBP5, PGC, PMEPA1*). In accordance, results showcased that 42.8% of PDXs progressed with downregulated AR and the analyzed AR downstream targets (**S1 Fig**).

### Metabolomic shifts in PCa response to castration

With the goal of understanding the metabolomic changes associated with PCa progression under ADT and identify the putative metabolites fueling CRPC, we used metabolomics techniques and bioinformatics tools. Briefly, tumor samples obtained from Control, ERC, and Relapse groups were subjected to UPLC-QTOF-MS/MS. The relative abundance of metabolites in Control, ERC, and Relapse tumors are depicted in the heatmap (**Fig 3 A**). 324 named metabolites were profiled (**Fig 3 B**). The comparison among groups using Welch’s Two-sample *t-*Test identified the following changes: 84 metabolites exhibited significant differential abundance (|Log2 fold change| > 1, P value < 0.05) when comparing ERC *vs*. Control tumors (22 lower and 62 higher); 66 in Relapse *vs*. Control tumors (26 lower and 40 higher); and 21 in Relapse *vs*. ERC tumors (16 lower and 5 higher) (**Fig 3 B**).

**Fig 3.**
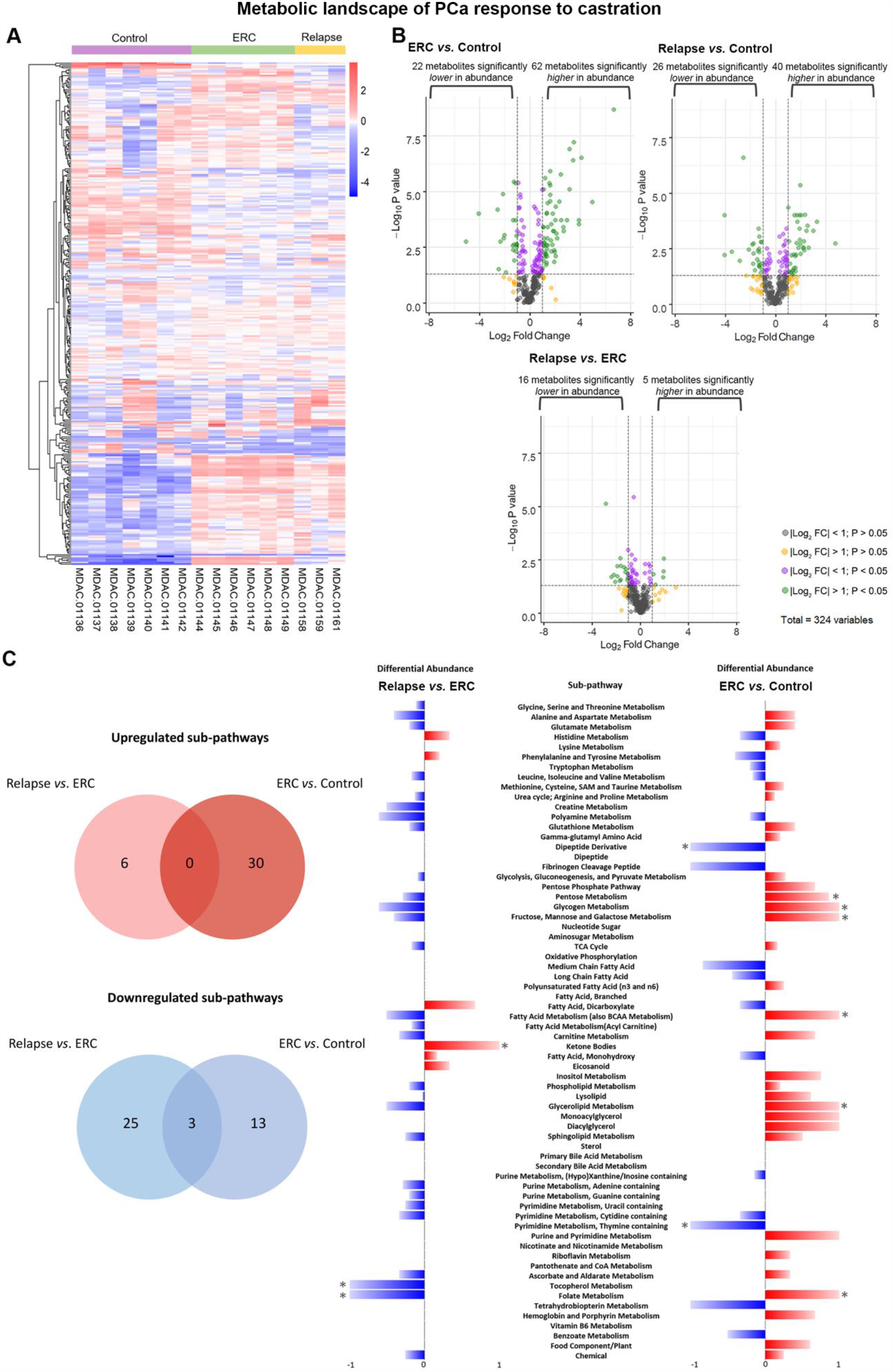
Metabolic shift of MDA PCa 183 tumors in progression to castration. (A) Heatmap depicting metabolites significantly higher or lower in Control (n=7), ERC (n=6) and Relapse (n=3) tumors. Raw counts were rescaled to set the median equal to 1 and expressed as imputed normalized counts for each metabolite (ScaledImpData). Log of the ScaledImpData was plotted. Relative abundance was color labeled (red to blue). (B) Volcano plots depict the 324 metabolites profiled. Significant differential abundance (P value < 0.05, |Log_2_ fold change| > 1) of higher and lower metabolites is represented for each comparison as green dots. Welch’s Two-Sample *t-*Test was used to calculate statistical significance. NS: not significant; Log_2_FC: log_2_ fold change. Grey dots: |log_2_ fold change|< 1 and P value > 0.05; yellow dots: |log_2_ fold change| > 1 and P value > 0.05; purple dots: |log_2_ fold change| < 1 and P value < 0.05; green dots: |log_2_ fold change| > 1 and P < 0.05. (C) KEGG pathway-based analysis of metabolic changes in Relapse *vs*. ERC and ERC *vs*. Control groups. Venn diagrams depict number of commonly and differentially up or downregulated sub-pathways in the comparisons. The differential abundance (DA) was defined as (No. of metabolites increased – No. of metabolites decreased) / No. of measured metabolites in pathway. Red horizontal bars indicate positive and blue horizontal bars indicate negative significant alterations in DA. * indicates enriched categories with a proportion of altered metabolites above 0.8 and a |Log2 fold change| > 1 of those metabolites.

Subsequently, we investigated the pathways associated with the above-mentioned metabolic profiles using KEGG pathway-based analysis. For these analyses we focused our attention on comparing Relapse *vs*. ERC and ERC *vs*. Control. In the first comparison, the tumor-bearing mice of both groups were castrated, but ERC tumors were not growing. Therefore, metabolites that increase in relapsed tumors (as compared with ERC) reflect changes associated with progression under ADT. In the second comparison, ERC *vs*. Control, differences in metabolite abundance reflect adjustment of ERC tumors to castration. Differential abundance (DA) scores were calculated (19), capturing the tendency for metabolites in a pathway to be increased or decreased for each comparison (**Fig 3 C, S2 Fig**). Among the 66 metabolic pathways captured in our profiling, 6 were up-regulated (red) and 28 were down-regulated (blue) for the Relapse *vs*. ERC comparison; and 30 were up-regulated and 16 were down-regulated for the ERC *vs*. Control comparison (**Figure 3 C**). Next, we plotted the proportion of metabolites in the sub-pathways that changed significantly for each comparison (x-axis), *vs*. the average log_2_(fold changes) of these metabolites (y-axis) (**Fig 4 A, S2 Fig**). For the ERC *vs*. Control comparison, the most significant elevated pathways were: glycogen, fructose, mannose & galactose, pentose, fatty acid, folate and glycerolipid metabolism; whereas pyrimidine (thymine containing) and dipeptide derivative were down-regulated pathways (**Fig 4 A**, left panel). Interestingly, the most significant pathway elevated in Relapse *vs*. ERC was involved in KB metabolism, whereas the decreased pathways were involved in tocopherol and folate metabolism (**Fig 4 A**, right panel). In particular, 3-hydroxybutyrate in the KB metabolism category had a >2 fold induction in Relapse *vs*. ERC tumors. On the other hand, alpha-tocopherol and 5MeTHF (5-methyltetrahydrofolate) were significantly downregulated in this comparison (**S3 Fig**).

**Fig 4.**
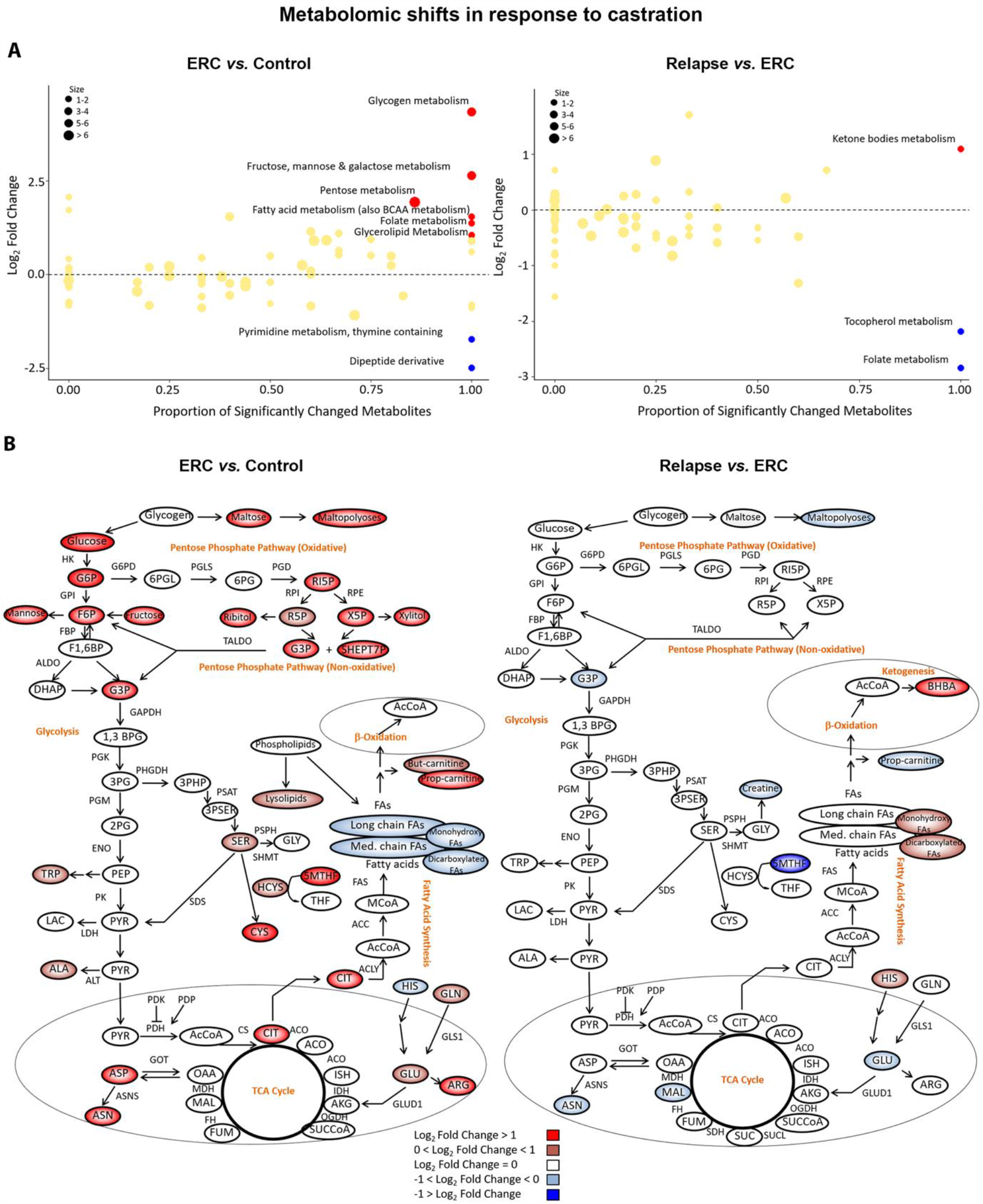
KEGG pathway-based analysis of metabolic changes upon comparison of Relapse vs. ERC and ERC vs. Control groups. (A) Scatter plots depicting altered KEGG pathways according to the metabolites increased and decreased in each comparison using Welch’s Two-sample *t*-Test. The x-axis indicates the proportion of metabolites in a pathway that are significantly changed (both increased and decreased) in the comparison. The y-axis plots the average log_2_ fold change of these metabolites. A proportion of significantly changed metabolites of 0.8 and a |Log2 fold change| > 1 were set as thresholds for pathways enrichment. (B) Metabolic changes of central carbon metabolism in ERC (n=6) *vs*. Control (n=7) or Relapse (n=3) *vs*. Control (n=7) comparisons. Metabolites are labeled as color-coded ovals (red: upregulated; blue: downregulated). Color corresponds to the log_2_ fold changes within a comparison. Enzymes for individual chemical reactions are denoted next to the arrows connecting two metabolites. *G6P*, glucose 6-phosphate; *F6P*, fructose 6-phosphate; *F1,6BP*, fructose 1, 6-bisphosphate; *DHAP*, dihydroxyacetone phosphate; *G3P*, glyceraldehyde 3-phosphate; *1,3BPG*, 1,3-bisphosphoglycerate; *3PG*, 3-phosphoglycerate; *2PG*, 2-phosphoglycerate; *PEP*, phosphoenolpyruvate; *TRP*, tryptophan; *PYR*, pyruvate; *LAC*, lactate; *ALA*, alanine; *3PHP*, 3-phosphohydroxypyruvate; *3PSER*, 3-phosphoserine; *SER*, serine; *AcCoA*, acetyl-CoA; *ISC*, isocitrate; *CIT*, citrate; *ACO*, cis-aconitate; *AKG*, alpha-ketoglutarate; *SUCCoA*, succinyl-CoA; *SUC*, succinate; *FUM*, fumarate; *MAL*, malate; *OAA*, oxaloacetate; *GLN*, glutamine; *GLU*, glutamate; *CYS*, cysteine; GLY, glycine; *MCoA*, malonyl-CoA; *HCYS*, homocysteine; *6PGL*, 6-phosphogluconolactone; *6PG*, 6-phosphogluconate; *R5P*, ribose 5-phosphate; *RI5P*, ribulose 5-phospate; *X5P*, xylulose 5-phosphate; *ASP*, aspartate; *ASN*, asparagine; *ARG*, arginine; *HIS*, histidine; *5MTHF*, levomefolic acid; *THF*, tetrahydrofolate; *BHBA*, β-hydoxybutirate; *SHEPT7P*, sedoheptulose-7-phosphate. ERC: early response to castration.

To better understand the metabolic alterations that occur upon relapse, we constructed a metabolic map detailing the shift in abundance of central carbon metabolites from Control *to* ERC and from ERC *to* Relapse (**Fig 4 B**). In the case of Relapse *vs*. ERC, we observed that the only increased metabolites were β-hydroxybutyrate (BHBA), decarboxylated FA (tetradecanedioate and hexadecanedioate), monohydroxy FA (13-HODE + 9-HODE) and histidine (**Figure 4 B**, right panel). In this comparison, it is noteworthy to mention the reduction in abundance of metabolites in the upper glycolysis and pentose pathways (PPP), as opposed to ERC *vs*. Control, where we observed a significant increase in metabolites involved in glycogen, pentose, mannose, fructose and galactose, metabolism (**Fig 4 B**, left panel). ERC activation of metabolites in upper glycolysis, including glucose 6-phosphate (G6P) and fructose 6-phosphate (F6P) (> 2-fold increases in abundance), suggests increased glucose uptake. Additionally, the shift toward the PPP observed in ERC *vs*. Control suggests increased shunting into the PPP to produce ribose 5-phosphate (R5P) and NADPH. Lysolipids are also increased to a lesser extent with concomitant reduction in FA, thus probably diverting energy production for biosynthesis of membranes and storage, favoring tumor dormancy (**Fig 4 B**, left panel).

### Dysregulated ketogenic/ketolytic enzymes in tumor-bearing PDX and human donor during PCa progression

Our previously described findings, showcase KB as one of the main metabolites highly increased during PCa growth under ADT. Therefore, we sought to evaluate enzymes involved in ketogenesis/ketolysis, namely ACAT1, 3-Hydroxybutyrate Dehydrogenase 1 (BDH1) and 3-oxoacid CoA-transferase 1 (OXCT1). ACAT1, BDH1 are reversible enzymes of the ketogenic/ketolytic pathway (**Fig 5 A**), allowing either the formation of KB or conversion to acetyl-CoA. However, OXCT1 catalyzes the irreversible reaction that transforms succinyl-CoA to acetoacetyl-CoA in most extrahepatic tissues (20). In line with the metabolomics data, we confirmed by immunohistochemical analysis the increased expression of the ketogenic enzymes (BDH1 and ACAT1) in Relapse *vs*. ERC (**Fig 5 B**). In the same comparison, we also observed increased expression of the extrahepatic enzyme OXCT1, which only participates in ketolysis (**Fig 5 B**). Hence, the increased levels of OXCT1 suggest utilization of KB as fuel by relapsed tumors. Most importantly, when we studied the expression of ACAT1, OXCT1, and BDH1 in the human donor tumor of MDA PCa 183, we detected increased expression of these enzymes in PCa progressing to ADT as compared with the untreated tumor of the same patient (**Fig 5 C**). These results support the concept that the changes observed in our PDX were predictive of the same changes that we identified in the human donor. Together, these data reflect the metabolic switch that accompanies relapse, where KB appear as the main energetic fuel driving PCa progression. This tumor cell energetic plasticity may denote an unseen landmark for early relapse in PCa.

**Fig 5.**
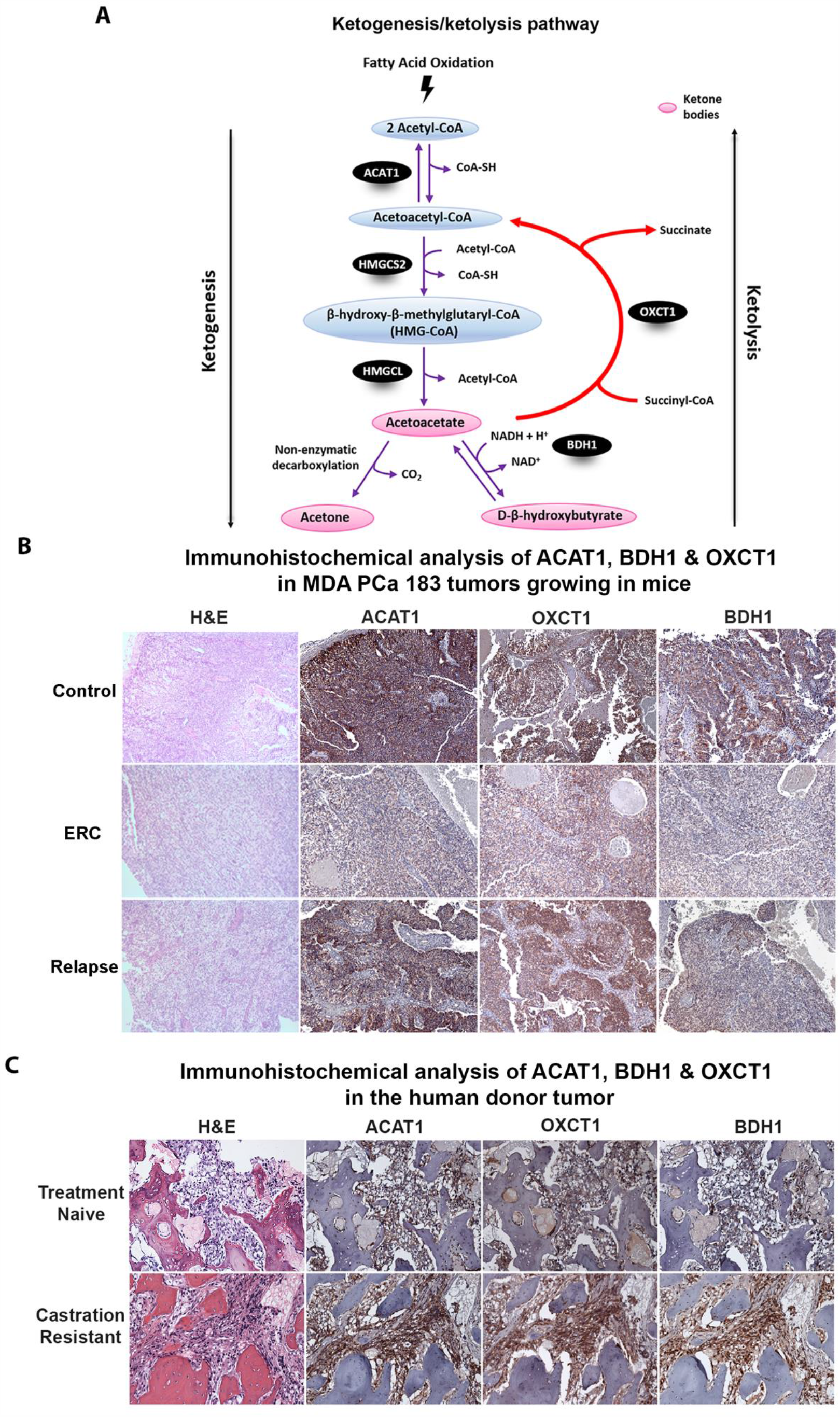
Upregulation of ketogenic/ketolytic enzymes in preclinical and clinical progression of PCa. (A) Schematic representation of ketogenesis/ketolysis pathway. The three ketone bodies are marked within a pink oval. Enzymes are depicted in black ovals and the other named metabolites in blue ovals. (B) Representative photomicrograph images of sections of MDA PCa 183 tumors growing in mice (ERC [n=6] and Relapse [n=3]). Magnification 100X. (C) Representative photomicrograph images of sections of samples of the corresponding human donor of MDA PCa 183 before ADT (treatment naive) and after progression (castration resistant). Magnification 200X. Samples were stained with H&E and immunostained for ACAT1, OXCT1 and BDH1. ERC: early response to castration; H&E: Hematoxylin and eosin; ACAT1: acetyl-CoA acetyltransferase; OXCT1: 3-oxoacid CoA-transferase 1; BDH1: 3-Hydroxybutyrate Dehydrogenase 1; HMGCS2: hydroxy-methylglutaryl-CoA synthase 2; HMGCL: hydroxy-methylglutaryl-CoA lyase.

### Analysis of ketogenic/ketolytic enzymes as risk predictors of clinical outcome in CRPC

As mentioned above, ACAT1, OXCT1 and BDH1 are critical in ketone metabolism. To further ascertain the implications of these factors in CRPC, we mined the Ross-Adams dataset (GSE70770, n=204). In-depth analysis of PCa patients that had undergone radical prostatectomy, revealed significant higher gene expression profiles for *ACAT1* (P=0.0006) and *OXCT1* (P=0.0009), in PCa patients that biochemically relapsed (BCR) within a 5-year follow-up period, as compared with patients that did not relapse in the same time-period (No BCR) (**Fig 6 A**). However, *BDH1* expression was reduced in BCR patients who relapsed later on (P=0.0002). Given that in our preclinical studies, relapsed tumors did not express ERG and AR expression was low (**Fig 2 B**), we also assessed *ERG* and *AR* expression in this dataset. Results indicated that BCR patients had reduced *ERG* levels (P=0.0009) and no significant changes were found for AR (**Fig 6 A**). In an independent analysis, when taking into consideration only patients that relapsed, those with low *AR* and *ERG*, presented high *ACAT1* and *OXCT1* expression (P=0.0013 and P=0.0034, respectively) and low *BDH1* (P=0.0048) (**Fig 6 B**). These results are in line ascertain our IHC observations, on the augmented expression of ACAT1 and OXCT1 in the preclinical model and the human donor upon relapse, but discrepancies arise for BDH1. Of note, BDH1 protein was found higher in our preclinical model and human donor upon relapse, while *BDH1* seemed downregulated at the transcriptional level in the Ross-Adams dataset. We cannot discard differential regulation at the translational or post-translational level that justify the accumulation of the protein which was detected by IHC (**Fig 5 B-C**) (21).

**Fig 6.**
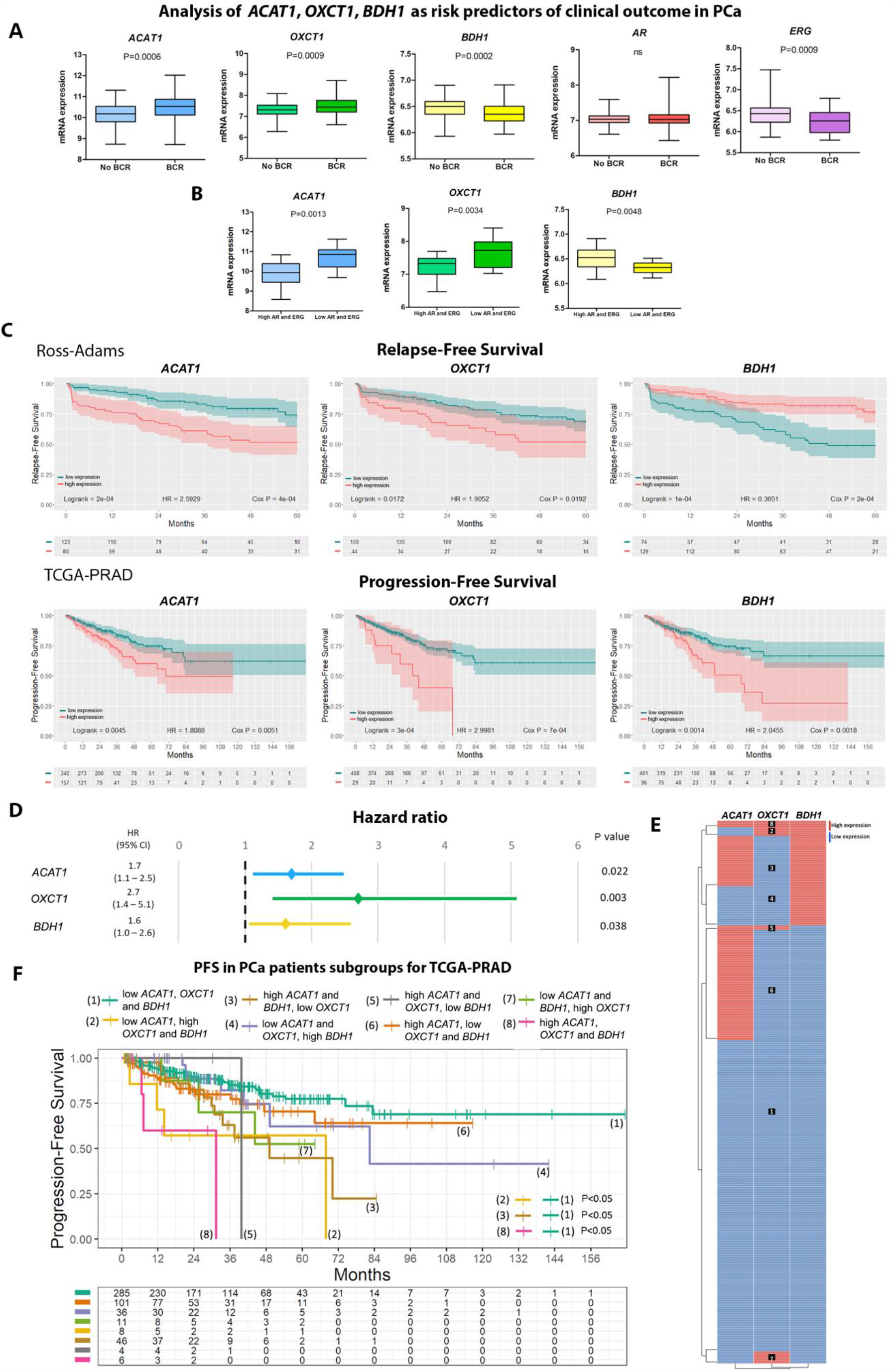
Bioinformatics analysis of ketogenic/ketolytic enzymes as risk predictors of clinical outcome in CRPC. (A) Gene expression analysis for *ACAT1, OXCT1, BDH1, AR* and *ERG* comparing PCa patients that underwent radical prostatectomy and biochemically relapsed (BCR; n=59) *vs*. those that did not relapse (No BCR; n=145) in the Ross-Adams dataset (GSE70770; n=204). Data are presented as box-and-whisker plots (min-max error bars). *t*-Test was used to assess statistical significance. (B) Gene expression analysis in PCa patients with BCR categorized by *ERG/AR* status (high expression n=16; low expression n=15). Data are presented as box-and-whisker plots (min-max error bars). *t*-Test was used to assess statistical significance. (C) Kaplan–Meier (KM) curves for relapse-free survival (RFS) (upper panels) and progression-free survival (PFS) (lower panels) in months for PCa patients with low (green) or high (red) *ACAT1, OXCT1* and *BDH1* expressions in the Ross-Adams and TCGA-PRAD (n=497) datasets, respectively. Log-rank test and Cox proportional hazard model regression were employed to assess statistical significance. (D) Multivariable Cox proportional hazard model regression analysis for *ACAT1, OXCT1* and *BDH1* presented as forest plots for PFS. (E) Heatmap depicting low (blue) or high (red) *ACAT1, OXCT1* and *BDH1* mRNA expression for PCa patients according to the TCGA-PRAD dataset. (F) KM curves for PFS in months for PCa patients subgroups with different expression levels of *ACAT1, OXCT1* or *BDH1* in TCGA-PRAD: (1) low *ACAT1, OXCT1* and *BDH1* expression (n=285); (2) low *ACAT1*, high *OXCT1* and *BDH1* expression (n=8); (3) high *ACAT1* and *BDH1* and low *OXCT1* expression (n=46), (4) high *BDH1* and low *ACAT1* and *OXCT1* expression (n=36), (5) high *ACAT1* and *OXCT1* and low *BDH1* expression (n=4); (6) high *ACAT1* and low *OXCT1* and *BDH1* expression (n=101); (7) high *OXCT1* and low *ACAT1* and *BDH1* expression (n=11) and (8) high *ACAT1, OXCT1* and *BDH1* expression (n=6). The table indicates the number of patients assessed every 12 months. Log-rank test was employed to assess statistical significance. All comparisons considered low expression patients as the reference group. HR: hazard ratio. Statistical significance: P < 0.05.

Next, we evaluated the relapse-free survival time (RFS) in PCa patients with BCR expressing high or low *ACAT1, OXCT1 and BDH1*. Results demonstrated significant association of high *ACAT1, OXCT1* expression with shorter RFS (**Fig 6 C**). The opposite was observed for *BDH1* (**Fig 6 C**). Interestingly, when performing the same analysis in the TCGA-PRAD dataset (n=497), high expression of *ACAT1, OXCT1* and *BDH1* was associated with shorter progression free survival (PFS) (**Fig 6 C**). Indeed, these results support the preclinical/clinical data about ACAT1 and OXCT1 expression as risk predictors upon relapse and evidence the discrepancies regarding BDH1.

To validate the potential of these molecules to improve risk stratification in PCa patients and in order to assess variable independence, multivariable analysis was performed between *ACAT1, OXCT1* and *BDH1* in the TCGA-PRAD dataset. Results showed that the three genes could be independent predictors of disease progression (P=0.022, P=0.003, P=0.038, respectively) (**Fig 6 D**). Next, we grouped PCa patients based on high or low *ACAT1, OXCT1* and *BDH1* gene expression levels. We categorized the patients into 8 groups according to the combinatorial expression of these three genes (**Fig 6 E**). We then performed a PFS analysis of these patient subgroups. Patients in groups 2, 3, and 8 had significantly decreased PFS compared with patients in group 1 (P<0.05 for all comparisons) (**Fig 6 F**). No significance was observed for the PFS for the rest of the comparisons (**Fig 6 F**). These results evidence the increased risk of progression for PCa patients with at least elevated expression of two of these genes.

## DISCUSSION

In this work we are reporting a metabolic shift towards ketogenesis/ketolysis that occurs in PCas that relapse after ADT. Furthermore, we have discovered that these metabolic alterations occur in cells that do not use the canonical AR signaling while progressing to ADT. Briefly we observed that relapsed tumors have an increase in KB content while tumor cells have reduced nuclear AR, increase of cytoplasmic AR and loss of ERG expression as compared with controls.

Reports evidence that metabolic changes are acquired during PCa progression and may underlie CRPC growth (22, 23). To further understand metabolic changes that occur during progression we performed a metabolomics analyses in a well-established PDXs that mimic donor response to ADT in a well-controlled study, which is difficult to achieve with clinical samples. The MDA PCa models have already provided unique insights into the biology of PCa (16), including the discovery of: distinct classes of chromosomal rearrangements in PCa cells (24); new therapeutic approaches for combination therapy targeting DNA damage response genes (25, 26); new biological roles of genes in PCa (27); new mechanisms underlying neuroendocrine differentiation (28); molecular alterations commonly found in aggressive variant PCa (29); fibroblast growth factor (FGF) axis in the pathogenesis of PCa bone metastasis (11, 14, 30, 31).

As mentioned earlier, the MDA PCa 183 PDX used to perform our studies, expresses wild-type AR, and has TMPRSS2/ERG rearrangement and ERG outlier expressions (15). However, when these tumors relapse, the change in AR subcellular localization and absence of ERG, indicate that these tumors progressed to castration without canonical AR signaling activation, reflecting progression with partial or complete loss of AR dependence.

Metabolomic analyses of MDA PCa 183 tumors after castration of tumor-bearing mice, showed a significant reduction in abundance of upper glycolysis and PPP metabolites, in Relapse *vs*. ERC as opposed to ERC *vs*. Control. ERC activation of metabolites in upper glycolysis, suggested increased glucose uptake. Additionally, the shift toward the PPP observed in ERC *vs*. Control suggests increased shunting into the PPP to produce R5P and NADP. These results depict that upon castration tumor cells activate upper glycolysis to survive and adapt.

Relapsed tumors had a significant increase in FA and KB compared with tumors at ERC. KB are high-energy mitochondrial fuels that can be converted back into acetyl-CoA and reutilized as an energy source (32, 33). Upon relapse, at least in our experimental model and human donor, PCa cells do not seem to use lactate to fuel their metabolism, as proposed by Warburg.

Since KB can be produced by β-oxidation of FA (34), which were also found to increase in relapsed tumors, we cannot rule out that FA could be the main source of energy implicated in CRPC progression. Under this scenario, KB could be a product of β-oxidation and a biomarker of CRPC progression. However, we confirmed that the expression of critical ketogenic/ketolytic enzymes - BDH1, OXCT1, and ACAT1- was significantly augmented after castration-resistant progression in MDA PCa 183 tumor. Furthermore, our discovery that these enzyme enhancements were also found in the human donor tissue after progressing to ADT add relevance to our findings as a mechanism of human PCa progression.

Expression of ACAT1, one of the key enzymes involved in the conversion of KB into acetyl-CoA, has been associated with aggressive PCa (22) and PCa biochemical recurrence following ADT (10). Thus, regardless of KB source of origin and because of the reported increase in ACAT1 in PCa and its association with relapse to ADT, KB appear as the high-energy fuel driving CRPC.

Most KB production occurs in the liver (34), however smaller amounts are generated in other tissues through altered expression of ketogenic enzymes (35, 36) or reversal of the ketolytic axis. Once the BHBA is incorporated by a tissue, it is converted back into acetoacetate by the same enzyme that generated it (BDH1). From there, the pathway of KB utilization diverges from the ketogenic synthetic pathway. Co-A is donated by succinyl-CoA to acetoacetate, to form acetoacetyl-CoA, a reaction catalyzed by OXCT1, bypassing the essentially irreversible reaction catalyzed by hydroxy-methylglutaryl-CoA synthase 2 (HMGCS2). As OXCT1 is an extrahepatic enzyme (34), these different enzymatic routes of synthesis and utilization of KB, prevent a BHBA useless cycle in the liver, allowing this metabolite to travel to other tissues as energy source. Thus, the acetoacetyl-CoA can then be converted back to two acetyl-CoA and incorporated into the Krebs cycle for oxidation and ATP formation (34). Of note, OXCT1 was significantly increased in relapsed MDA PCa 183 tumors and the human donor progressing on ADT therapy (castrate resistant). This result evidences a tilt in the balance of the ketogenic/ketolysis pathway towards acetyl-CoA formation, vital for amino-acids, ATP and FA synthesis and subsequently for tumor progression.

Interestingly, in our univariable time-to-recurrence analysis in the TCGA-PRAD dataset, a positive association of increased expression of *ACAT1, OXCT1* and *BDH1* with PCa progression was observed. When performing multivariable analyses, the three transcripts appear to behave independently when predicting risk of progression. Further, when regrouping PCa patients according to *ACAT1, OXCT1* and *BDH1* expression levels, results evidenced the increased risk of progression for PCa patients with co-occurrence for these elevated transcripts.

Reports have shown ACAT1 expression to predict recurrence in ERG-negative cases, whereas ERG-positive cases did not display any difference (10). However, no reference was provided as to whether the ERG-negative cases presented a *TMPRSS2*/*ERG* translocation that had bypassed the AR signaling or simply did not present the translocation at all. For this reason, ACAT1 appears as an exploitable target to assess its implications accompanying PCa relapse to ADT associated with low or negative AR expression.

ACAT1 was proposed as a druggable target for cancer therapy since in its active phosphorylated tetrameric form, catalyzes the conversion of two acetyl-CoA to acetoacetyl-CoA and CoA and also acts as an upstream acetyl transferase for the pyruvate dehydrogenase complex, a decisive point between two paths: glycolysis and oxidative phosphorylation (37). In many cancer types, active ACAT1 inhibits pyruvate dehydrogenase activity and deflects pyruvate to lactate production. However, our results demonstrated high expression of ACAT1 in Relapse *vs*. ERC tumors, with augmented KB but no increase in lactate production. Since KB appeared as the main route fueling energy during relapse to ADT, inhibition of ACAT1 may block this energy source rather than trigger lactate production in this type of tumor. Moreover, ACAT1 is common to both, the mevalonate pathway (biosynthesis of cholesterol, sterols, etc.) and KB production, hence targeting this enzyme appears to be promising for therapeutic intervention of CRPC.

Our work has identified critical metabolic changes in early relapse following ADT of a subpopulation of PCa that progress with low AR/ERG. These metabolic changes may serve as the foundation to identify early biomarkers of CRPC progression in a larger study.

## METHODS

### MDA PCa 183 PDX generation

At MD Anderson, we established a program to develop PCa PDXs (the MDA PCa PDX series) using clinically annotated PCa specimens with the goal of more comprehensively modeling the complexity of PCa. The PDX MDA PCa 183 was developed in our laboratory from a bone marrow aspirate of a human male with treatment naïve metastatic adenocarcinoma (15, 16, 18) as described previously (16), and propagated as subcutaneous xenografts in 6- to 8-week-old male CB17 SCID mice (Charles River Laboratories).

### MDA PCa 183 growth in intact and castrated mice

Studies were conducted as described elsewhere (16). Briefly, 6- to 8-week-old male CB17 SCID mice (n=20) were subcutaneously implanted with MDA PCa 183. Castration was performed in 13 of these mice when tumors reached a volume of 500 mm^3^ or higher (castrated mice). Tumor volume was monitored in intact and castrated mice. Tumors from intact mice were harvested when the volume reached 2000 mm^3^ (Control). Tumors from 6 of the castrated mice were harvested at 10 days of castration (early response to castration (8)). The remaining 7 continued to be monitored for tumor growth over time and the relapsed tumors were harvested. Relapsed tumors are those that continue to grow for two consecutive weeks after castration (Relapse). PSA was measured using PSA ELISA kit (American Qualex).

### Transcriptome analysis of CRPC MDA PCa PDXs

Samples were prepared as previously described (11). RNA was extracted from fresh frozen tissue of 37 CRPC MDA PCa PDXs (16, 18) at the Biospecimen Extraction Facility (MD Anderson), using the QIAGEN RNeasy kit. Stranded total RNA-sequencing and transcriptome analysis were performed as previously described (38), at the Advanced Technology Genomics Core (MD Anderson), using the NovaSeq 6000 SP flow cell with 100nt PE sequencing format. AR and AR downstream targets gene expression was analyzed using the pheatmap package in R (39).

### Non-targeted global metabolite profiling

Sample preparation and analysis was carried out at Metabolon, Inc. In brief, sample preparation involved protein precipitation and removal with methanol, shaking and centrifugation. The resulting extracts were divided into fractions for analysis on three independent platforms: ultra-high-performance liquid chromatography/tandem mass spectrometry (UHPLC/MS/MS) optimized for basic species, UHPLC/MS/MS optimized for acidic species, and GC/MS. The details of this platform have been described previously (40). Metabolites were identified by automated comparison of the ion features in the experimental samples to a reference library of chemical standard entries that included retention time, molecular weight (*m/z*), preferred adducts, and in-source fragments as well as associated MS spectra, and were curated by visual inspection for quality control using software developed at Metabolon (41). Values obtained were normalized in terms of raw area counts (OrigScale). Each biochemical in OrigScale was rescaled to set the median equal to 1 and expressed as imputed normalized counts for each biochemical (ScaledImpData).

### Immunohistochemistry

We performed immunohistochemistry (IHC) analyses of AR and ERG expression in mouse tissue specimens obtained from MDA PCa 183 tumors in Control (n=5), ERC (n=6) and Relapse (n=3) groups. We also performed IHC analyses of ACAT1, OXCT1, BDH1 in sections derived from these mouse specimens and in bone biopsies obtained from the human donor of MDA PCa 183 untreated and after relapse. Donor samples were obtained from the Prostate Tissue Bank, Department of Pathology, MD Anderson Cancer Center, Houston, TX, under an Institutional Review Board approved protocol. Bone specimens were decalcified and all samples formalin-fixed, paraffin-embedded as previously described (42). Tissue sections were stained with anti-AR (N-terminal) antibody (Dako), anti-ERG antibody (Biocare Medical), anti-ACAT1 antibody (Sigma), anti-BDH1 antibody (Sigma) and anti-OXCT1 antibody (Sigma) as described elsewhere (43).

### Animals

All practices involving laboratory animals were conducted under the approval of the Institutional Animal Care and Use Committee of The University of Texas MD Anderson Cancer Center, under the regulation of the Animal Welfare Committee (IACUC), and conform to the NIH Policy on Humane Care and Use of Laboratory Animals.

### Statistics

Welch’s two sample t-test was used to ascertain statistical significance. Heatmap was created in R with the pheatmap package (39) using the log of the ScaledImpData. Volcano plots and scatter plots were performed using EnhancedVolcano (44) and ggplots2 (45) packages in R, respectively. Differential abundance (DA) score was calculated by first applying a non-parametric differential abundance test (in this study, Welch’s Two-Sample t-Test) to all metabolites in a pathway. After determining which metabolites were significantly increased/decreased in each comparison, differential abundance score was defined as DA = (No. of metabolites increased – No. of metabolites decreased) / No. of measured metabolites in pathway. |Log_2_ fold change| > 1, proportion of significantly changed metabolites > 0.8 and P value < 0.05 were set as threshold for statistical interpretation (19).

## Bioinformatics analysis

### Information source and eligibility criteria (The Cancer Genome Atlas (TCGA) (46))

To study the impact of the expression of the selected genes on the progression-free survival (PFS) of PCa patients, we used the Xena platform (47) to access the dataset from the Prostate Adenocarcinoma Project of The Cancer Genome Atlas (TCGA-PRAD) (http://cancergenome.nih.gov/). TCGA-PRAD has gene expression data from 497 prostate tumor samples and normal adjacent tissue (last access: June 2020), measured by massively parallel sequencing (llumina HiSeq).

### Information source and eligibility criteria (GEO: Gene Expression Omnibus)

To study the impact of gene expression levels on relapse-free survival (RFS) in PCa patients, we selected the Ross-Adams 2015 dataset (GSE70770) (48). GPL10558 series, according to the following criteria: (1) the study included gene expression and clinical data for each patient with ≥ 5 years of follow-up; (2) the study consisted of ≥ 60 samples; and (3) the study was published and available in public repositories. Tumor sample expression of 31,000 transcripts was measured by 47,000 probes using the Illumina HumanHT-12 V4.0 platform This PCa patient cohort included 204 samples from men who had undergone radical prostatectomy and clinical follow-up up to 8 years, including relapse information. Biochemical relapse was defined according to the European Guidelines as a prostate specific antigen (PSA) persistent rise above 0.2 ng/mL.

### Statistics

For *AR/ERG* gene expression categorization in patients that biochemically relapsed within five years from the Ross-Adams dataset (GSE70770), we established the median of each gene expression level as the cutoff point (> median: high expression; < median: low expression). *t*-Test was used to assess the statistical significance using GraphPad Prism software (La Jolla, CA, USA). Survminer R package (49) was used to explore patients’ RFS and PFS, and to generate Kaplan–Meier (KM) curves. To find the cutoff value to stratify patients into two groups based on the gene expression levels for time to event analysis, we used the minimal P value approach from the Cutoff Finder tool (50). For univariable and multivariable analyses, the log-rank test and Cox proportional hazard model regression were employed. The hazard ratio (HR) indicates the probability of an event in a patient with high gene expression with respect to one with low gene expression, considering HR=1 for the latter case. *t*-Test was used to assess the statistical significance set as P < 0.05.

## Data Availability

Not applicable

## Acknowledgments

We thank Jordan T. Pietz for the scientific illustrations, the Biospecimen Extraction Resource of MD Anderson Cancer Center for the sample processing and RNA extraction, and the Advanced Technology Genomics Core of MD Anderson [CA016672(ATGC)].

## Author contributions

E.V., G.G. and N.N. designed the experimental approach; E.L., J.Y., A.P., P.D.A.S., and A.G.H. performed laboratory-based studies; E.L., J.B., P.S., V.A.A., N.A., S.L.-V., J.C., E.V., G.G. and N.N. evaluated all the results; E.L., J.B., P.S., E.V., G.G. and N.N. wrote the manuscript, and prepared the figures; E.L., J.B., P.S., J.C., E.V., G.G. and N.N. designed and performed bioinformatics and statistical analysis; J.A., C.L. and E.E. provided clinical oncological expertise and conceptual insight; E.V., G.G. and N.N. provided overall project design, management, and supervision.

