## Supplemental information for "Prostate cancer castrate resistant progression usage of non-canonical androgen receptor signaling and ketone body fuel"

SUPPORTING INFORMATION

Supplementary Figure 1

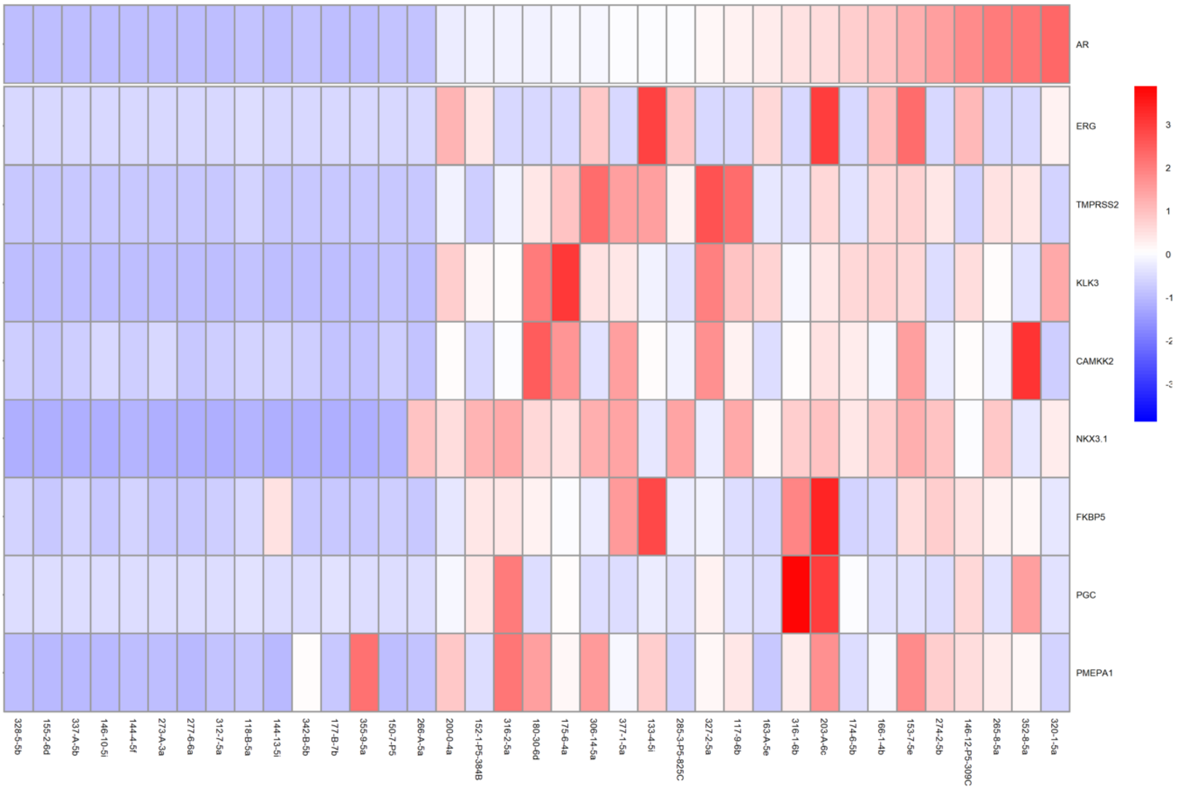


**S1 Fig. *AR* and AR downstream targets' gene expression in CRPC MDA PCa PDXs.** Analysis of RNAseq data from 37 CRPC MDA PCa PDXs showcased in a heatmap depicting *AR* and AR downstream targets' (*ERG*, *TMPRSS2*, *KLK3*, *CAMKK2*, *NKX3.1*, *FKBP5*, *PGC*, *PMEPA1*) gene expression. CRPC PDXs identifiers are shown in the x-axis. Red, white, and blue represent greater, intermediate, and lower gene expression levels, respectively. Expression values are expressed as z-score.

Supplementary Figure 2

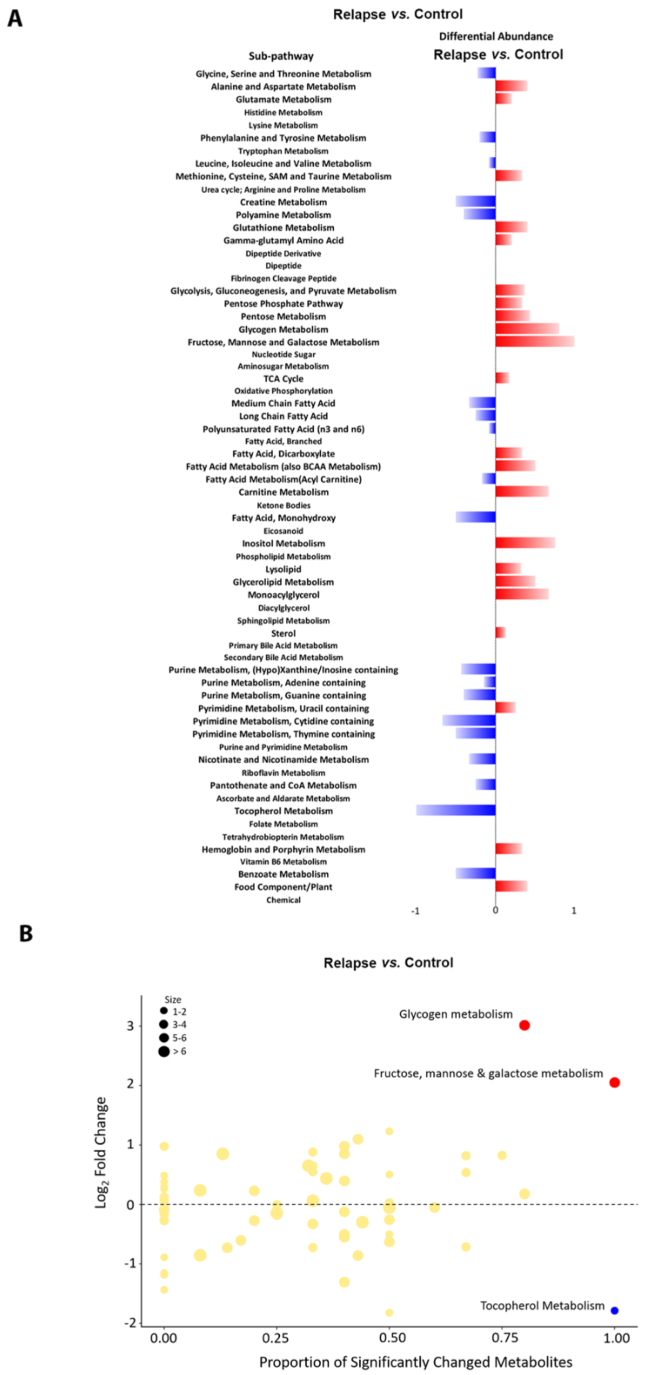


**S2 Fig. Metabolic shift of MDA PCa 183 Relapse *vs*. Control tumors tumors.** (A) KEGG pathway-based analysis of metabolic changes in Relapse *vs*. Control. The differential abundance (DA) was defined as (No. of metabolites increased – No. of metabolites decreased) / No. of measured metabolites in pathway. Red horizontal bars indicate positive and blue horizontal bars indicate negative significant alterations in DA. (B) Scatter plots depicting altered KEGG pathways according to the metabolites increased and decreased in Relapse *vs*. Control using Welch's Two-sample t-Test. The x-axis indicates the proportion of metabolites in a pathway that are significantly changed (both increased and decreased) in the comparison. The y-axis plots the average log2 fold change of these metabolites. A proportion of significantly changed metabolites of 0.8 and a |Log2 fold change| > 1 were set as thresholds for pathways enrichment.

Supplementary Figure 3

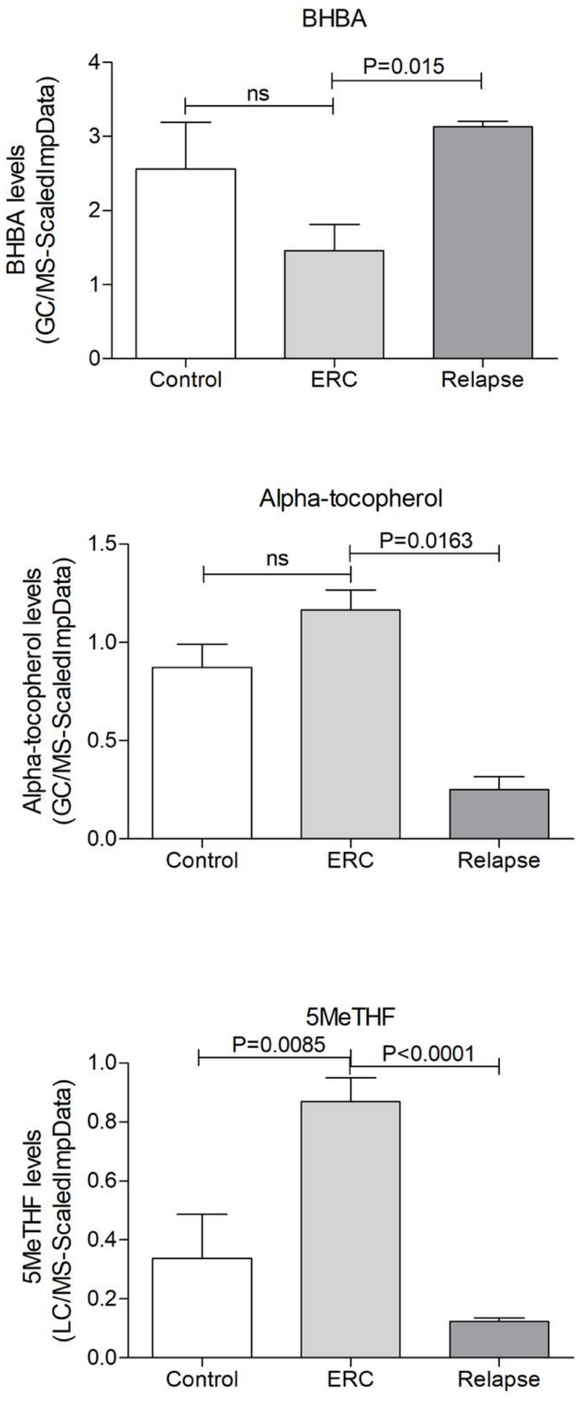


**S3 Fig. Relative abundance of metabolites altered in Relapse *vs.* ERC**. Bar plots of the abundance of BHBA (3-hydroxybutyrate), alpha-tocopherol and 5MeTHF (5-methyltetrahydrofolate) measured by GC/MS or LC/MS in Control (n=7), ERC (n=6) and Relapse (n=3) mice. Statistical significance was assessed using Welch's Two-sample t-Test between each comparison and was set at $P < 0.05$. Values represent the ScaleImpData for each metabolite. Error bars: SEM.
